# Colonic mucosal associated invariant T cells in Crohn’s disease have a diverse and non-public T cell receptor beta chain repertoire

**DOI:** 10.1101/2023.05.05.23289565

**Authors:** Andrew J Konecny, Donna M Shows, James D Lord

**Author notes:** Corresponding Author: James D Lord, MD, PhD Benaroya Research Institute, 1201 Ninth Ave., Seattle, WA 98101, U.S.A.

## Abstract

**Objectives:** Mucosal-Associated Invariant T (MAIT) cells are T cells with a semi-invariant T cell receptor (TCR), recognizing riboflavin precursors presented by a non-polymorphic MR1 molecule. As these precursors are produced by the gut microbiome, we characterized the frequency, phenotype and clonality of MAIT cells in human colons with and without Crohn’s disease (CD).

**Methods:** The transcriptome of MAIT cells sorted from blood and intestinal lamina propria cells from colectomy recipients were compared with other CD8^+^ T cells. Colon biopsies from an additional ten CD patients and ten healthy controls (HC) were analyzed by flow cytometry. TCR genes were sequenced from individual MAIT cells from these biopsies and compared with those of MAIT cells from autologous blood.

**Results:** MAIT cells in the blood and colon showed a transcriptome distinct from other CD8 T cells, with more expression of the IL-23 receptor. MAIT cells were enriched in the colons of CD patients, with less NKG2D in inflamed versus uninflamed segments. Regardless of disease, most MAIT cells expressed integrin αEβ7 in the colon but not in the blood, where they were enriched for α4β7 expression. TCR sequencing revealed heterogeneity in the colon and blood, with few public sequences associated with cohorts.

**Conclusion:** MAIT cells are enriched in the colons of CD patients and disproportionately express molecules (IL-23R, integrin α4β7) targeted by CD therapeutics, to suggest a pathogenic role for them in CD. Public TCR sequences were neither common nor sufficiently restricted to a cohort to suggest protective or pathogenic antigen-specificities.

## INTRODUCTION

Mucosal-Associated Invariant T (MAIT) cells are a predominantly CD8^+^[1, 2] sub-population of αβ T cells that were first identified for their invariant use of T cell receptor (TCR) alpha genes TRAV1-2 (TCRVα7.2) and TRAJ33.[3] MAIT cells usually represent about 1-10% of circulating CD3^+^ lymphocytes in humans, and are enriched in mucosal tissues of the lung, liver, genitourinary tract, and gastrointestinal tract.[4],[5] MAIT cell TCRs do not recognize MHC-bound peptides, but rather riboflavin precursors[6] presented by the genetically non-polymorphic major histocompatibility complex class-1 related protein 1 (MR1).[7] These riboflavin precursors are not synthesized by mammalian cells but are instead produced by certain bacteria and yeast, many of which are common to the commensal flora of the gastrointestinal tract. In this way MAIT cells can monitor a diverse array of microbiota through their metabolic products[6]..

Perhaps as a consequence, MAIT cells are central to early antimicrobial immunity at mucosal barriers and specifically have been shown to mount an immune response to *Mycobacterium tuberculosis* independent of prior infection.[8] Circulating MAIT cells are also reduced in individuals with active *Mycobacterium tuberculous*,[8–10] *Vibrio cholerae* O1,[11] as well as *Salmonella typhi* infections,[12] viral infections,[1, 13–16] and severe bacterial infections resulting in sepsis.[17] It is hypothesized that MAIT cell depletion from the blood is a direct result of their recruitment to tissues where they combat active infections.[8, 18]

A similar decrease in peripheral MAIT cells is found in patients with Multiple Sclerosis (MS),[19] systemic lupus erythematosus (SLE) and rheumatoid arthritis (RA),[20] Sjögren’s syndrome,[21] Type 1[22] and 2[23, 24] Diabetes, celiac disease,[25] ulcerative colitis (UC) and Crohn’s disease (CD).[26–29] A reciprocal increase of MAIT cells was observed in the synovial fluid of RA patients[20] and accumulation of MAIT cells in central nervous system lesions and peripheral nerves of MS patients suggesting that MAIT cells are recruited to inflamed tissues in these settings of autoimmunity.[19, 30–33] However, histological reports differ as to whether MAIT cells are increased[28, 29] or decreased[26] in the bowel in CD. In two studies, MAIT cells in blood were reported to be more activated in CD, with more Ki67, BTLA, and NKG2D expression [28] as well as more expression of the cytokines TNF and IL-17A,[29] suggesting they are also qualitatively more pro-inflammatory in CD. However, in another study, MAIT cells expressed more activated caspases in CD, suggesting they are more prone to apoptosis,[26] in which case their elimination could be pathogenic if they normally play an immunoregulatory role, or limit the activity of microbes that may contribute to disease.

CD is an idiopathic inflammatory bowel disease (IBD) whose pathophysiology shares certain features with the immune response to tuberculosis, such as the formation of granulomas in tissue[34] and vulnerability to TNF blockade.[35] Indeed, this has led some to hypothesize that an unknown mycobacterial infection could underlie CD,[36] as genetic susceptibility factors for CD show striking overlap with those for mycobacterial infections in genome-wide association studies.[37] Given their role in mycobacterial infections, MAIT cells may likewise be central to CD immunopathology.

MAIT cells share many properties with Th17 cells, an IL-17A-producing CD4^+^ population of T cells that has been much more extensively studied than MAIT cells in IBD,[38, 39] and has been implicated in the pathogenesis of multiple autoimmune diseases and disease models.[40–42] Like Th17’s, MAIT cells are described as “effector/memory” being CD45RA^-^, CCR7^-^, and CD62_low._[8, 43] In addition, they both express the transcription factor RAR-related orphan receptor gamma (RORγt),[43, 44] the natural killer (NK) cell markers CD161 and CD26;[8, 45] tissue homing receptors CCR2, CCR5, CCR6, CXCR6, and CCR9;[1, 8, 46–49] cytokine/effector molecules TNF, IL-17A, and IL-22,[8, 10, 43] and receptors for such cytokines as IL-7 and 23.[1, 50–53] Th17 cells require the cytokine IL-23 for differentiation and survival,[6, 54, 55] and CD has been strongly associated with genetic polymorphisms of the IL-23 receptor,[56] including a rare coding mutation that protects people from developing CD.[57] Furthermore, multiple trials of humanized antibodies blocking IL-23 have successfully treated CD,[58–61] making IL-23 blockade a major FDA-approved treatment strategy for CD. While the dependence of Th17 cells upon IL-23 for survival suggests that these cells are thus central to CD, there has been little to no data demonstrating that Th17 cells are depleted by anti-IL-23 therapy,[62, 63] despite the latter having been in clinical use for psoriasis for more than a decade.[64, 65] However, MAIT cells have also been shown to respond to IL-23, which augments their antigen-dependent production of IL-17A.[66–68] Thus, both the genetic and pharmacological data implicating IL-23 in CD pathogenesis may just as well support a role for MAIT cells as Th17’s.

We describe a more detailed analysis of MAIT cells in CD than has previously been published, including both flow cytometry and full genome transcriptome profiling of these cells in the intestinal mucosa. Furthermore, we describe paired TCR alpha and beta chain gene sequences in individual MAIT cells from colon biopsies, paired with TCR beta sequences from autologous blood MAIT cells, to report on the clonality, diversity, and potentially public nature of these semi-invariant cells in the GI tract in health and disease.

## MATERIALS AND METHODS

### Patient specimens

All human cells were obtained from a biorepository at the Benaroya Research Institute, to which deidentified, consenting donors had previously contributed under a protocol authorized by our institutional review board (IRB), in compliance with the Declaration of Helsinki. For transcriptome profiling of intestinal MAIT and CD8^+^ T cells, frozen vials of colon lamina propria mononuclear cells (LPMC) were obtained, which had previously come from the surgical resections of six CD patients, six UC patients, and six patients without IBD as the indication for surgery. The resected tissue was inflamed in four of the CD patients, and not inflamed in one CD patient and all six patients without IBD. In the sixth CD patient and all six UC patients, both inflamed and uninflamed samples of colon were both available to be analyzed independently.

For flow cytometry analyses of colon MAIT cells, as well as single cell sorting for TCR sequencing, vials containing frozen colonoscopic biopsies were obtained from ten healthy screening colonoscopy recipients and ten CD patients. Biopsies from inflamed and uninflamed colon were retrieved in separate vials for each of the CD patients. Frozen vials of peripheral blood mononuclear cells (PBMC) were also available for analysis from eight of subjects in each of these cohorts. Clinical and demographic details about these donors are presented in Supplementary table 1.

### Specimen processing

Surgically resected colon specimens were processed for homogenized lamina propria (LP) cells and frozen as previously described.[69] Colonoscopic jumbo forceps biopsies were placed into bovine calf serum (BCS) at the bedside and promptly transported to the laboratory on ice where an equal amount of 14% DMSO in BCS was added for a final concentration of 7% DMSO for cryopreservation. Biopsies were frozen slowly in freezing jars (−70° C) following transfer into liquid nitrogen storage (−120° C) until use. PBMC were isolated using Lymphoprep™ (Axis-Shield, Oslo, Norway) density gradient, suspended in 7% DMSO in BCS, and frozen in the same manner.

### MAIT FACS and RNAseq

Homogenized LP and PBMC were thawed and stained extracellularly with UV Green or Blue Live/Dead Dye (Invitrogen, Waltham, MA, USA) and antibodies to CD3 (clone SK7, BioLegend, San Diego, CA, USA), CD4 (clone RPA-T4, BioLegend), CD8α (clone RPA-T8, eBioscience, San Diego, CA, USA), CD45RA (clone HI-100, BioLegend), CD103 (clone Ber-ACT8, BioLegend), CD161 (clone HP-3G10, BioLegend), CD314 (NKG2D, clone 1D11, BioLegend), TCRγδ (clone B1, BioLegend), and TCR Vα7.2 (clone 3C10, Biolegend). For PBMC, antibodies to integrin α4 (CD49d, clone 9F10, BD Biosciences, NJ, USA) and β7 (clone FIB504, eBioscience) and a fluorophore-conjugated version of the anti-integrin α4β7 biopharmaceutical vedolizumab (Takeda, Tokyo, Japan) were also included. Cells were then immediately sorted live using a BD FACSAria Fusion. MAIT cells were defined by singlets, lymphocytes, live (Green Live/Dead negative), CD3^+^, CD4^-^, TCRγδ^-^, and TCR Vα7.2^+^, CD161^high^ and compared to CD8 effector T cells defined as singlets, lymphocytes, live (Green Live/Dead negative), CD3^+^, CD4^-^, CD8^+^, and CD45RA^-^, from which MAIT cells, above, had already been excluded. One thousand CD8 effector T cells and MAIT cells were sorted into lysis buffer and processed for bulk RNA sequencing using the SMARTseq v4 platform (Takara Bio, Kusatsu, Japan). Flow cytometry data was analyzed with FlowJo Software (BD Biosciences). Representative gating strategies are shown in Supplementary figures 1-3.

### TCR sequencing

Intact biopsies from HC and CD patients (with paired inflamed and non-inflamed colon specimens from the latter only) were thawed and enzymatically digested in RPMI supplemented with HEPES, PSG, BCS, MgCl_2_, CaCl_2_, Collagenase Type 1, and 0.011% DNase for 30 minutes at 37° C. Cells were then pelleted, resuspended, and mechanically digested by passing the suspension through an 8-gauge needle and filtered through a 100-μum screen. Cells were again pelleted, stained as above, and immediately sorted using the BD FACS Fusion. Live single MAIT cells were sorted into a 96-well plate and their TCR alpha and beta chain amplified using a nested-PCR protocol.[70] PCR products were Sanger sequenced by a commercial third party (GeneWiz; now Azenta Life Sciences, Chelmsford, MA) using primers specific to the common region of the TCR alpha or beta gene, and TCR alignment was performed using the IMGT HighV-quest tool. For blood TCR analyses, up to 50,000 MAIT cells were sorted by FACS as above from thawed PBMC. DNA extracted from sorted MAIT cells was then sent for comprehensive TCR beta chain sequencing by a commercial third party (Adaptive Biotechnologies, Seattle, WA), with data thus generated retrieved from and analyzed on the latter’s proprietary online ImmunoSEQ Analyser software.

### Bulk RNA-seq experiments and analysis

Cells were sorted directly into lysis buffer from the SMART-Seq v4 Ultra Low Input RNA Kit for sequencing (Takara). Cells were then lysed, and cDNA was synthesized and amplified per the manufacture’s instruction. After amplification, sequencing libraries were constructed using the NexteraXT DNA sample preparation kit with unique dual indexes (Illumina, San Diego, CA) to generate Illumina-compatible barcoded libraries. Libraries were pooled and quantified using a Qubit Fluorometer (Life Technologies, Carlsbad, CA). Sequencing of pooled libraries was carried out on a NextSeq 2000 sequencer (Illumina) with paired-end 59-base reads, using a NextSeq P2 sequencing kit (Illumina) with a target depth of 5 million reads per sample.

Base calls were processed to FASTQs on BaseSpace (Illumina), and a base call quality-trimming step was applied to remove low-confidence base calls from the ends of reads. Reads were processed using workflows managed on the Galaxy platform. Reads were trimmed by 1 base at the 3′ end then trimmed from both ends until base calls had a minimum quality score of at least 30. Any remaining adapter sequence was removed as well. To align the trimmed reads, STAR aligner (v2.4.2a) was used with the GRCh38 reference genome and gene annotations from ensembl release 91. Gene counts were generated using HTSeq-count (v0.4.1). Quality metrics were compiled from PICARD (v1.134), FASTQC (v0.11.3), Samtools (v1.2), and HTSeq-count (v0.4.1).

A quality filter was applied to retain libraries in which the fraction of aligned reads examined compared to total FASTQ reads was > 70%, the median coefficient of variation of coverage was less than 0.85, and the library had at least 1 million reads. All analyzed samples passed these quality filters. Non-protein coding genes and genes expressed at less than 1 count per million in fewer than 10% of samples were filtered out. Expression counts were normalized using the TMM algorithm. For differential gene expression analysis, the linear models for microarray data (Limma) R package after Voom transformation was used; this approach either outperforms or is highly concordant with other published methods. Linear models were generated, and donor identity was included as a random effect. For differential gene expression comparisons, genes with a false discovery rate (FDR) of less than or equal to 0.05 and an absolute expression fold-change greater than or equal to 1 were considered differentially expressed.

### Statistical analysis

Analyses were preformed with Prism software (GraphPad). Comparisons between groups of flow cytometry data and other continuous variables did not presume a Gaussian distribution. Therefore, analysis of variance used a Kruskal-Wallis test for unpaired data and a Friedman test for paired comparisons. Two-way comparisons were performed using a Mann-Whitney *U*-test for unpaired comparisons and a Wilcoxon signed-rank test where data points could be paired (e.g. were from the same donor). Specific tests employed are specified in figures and/or their legends. Given the exploratory nature of these analyses, *P*-values were not adjusted for multiple comparisons.

## RESULTS

### Transcriptome analysis of MAIT cells in colon and blood

To compare the gene expression of MAIT (CD3^+^, CD4^-^, CD161^+^, TCRVα7.2^+^) cells to that of CD8^+^ effector (CD3^+^, CD4^-^, CD8a^+^, CD45RA^-^) T cells from which these MAIT cells were excluded, cDNA was analyzed by RNAseq from each of these two populations after they were sorted by FACS from patient specimens. Although MAIT cells in blood can also be identified with MR1 tetramers, these were not employed because we have found tetramers to poorly label T cells from collagenase-treated colon (data not shown). Thus, a small subpopulation of CD4^+^ MAIT cells could not be defined by CD161 and TCRVα7.2, due to CD161 also being expressed by Th17 cells which are enriched in the colon. All CD4^+^ T cells were therefore excluded from these and subsequent analyses of MAIT cells. Cells were sorted from the crypreserved PBMC of 6 CD patients, 6 UC patients, and 3 patients without IBD, all of whom were undergoing surgical resection of colon. Additionally, from among the above PBMC donors, these two T cell populations were sorted from cryopreserved LPMC isolated from the surgically resected colonic tissue of 1 non-IBD patient (who underwent sigmoid colon resection as part of a rectopexy procedure for rectal prolapse), 1 CD patient with inflamed colon, 1 CD patient with non-inflamed colon, 1 CD patient with both inflamed and non-inflamed colon in separate specimens, 1 UC patient with non-inflamed colon, and 3 UC patients with both inflamed and non-inflamed colon in separate specimens. Both CD4^-^ MAIT and CD8^+^ effector populations from all 27 of the above specimens produced good-quality libraries, resulting in 2.5 to 12 million reads each, with at least 70% gene alignment and a CV of coverage < 0.8 in all cases. Principal component (PC) analyses divided specimens according to location (blood versus colon) in PC1 and PC2 (Fig 1**a**), while dividing them by cell type (CD4^-^ MAIT vs effector CD8 T cells) in PC3 and PC4 (Fig 1**b**). These cohort sizes were obviously too small to make multivariate comparisons between diseases or the presence of inflammation, so data was pooled for all colon specimens in the subsequent analyses.

**Fig 1:**
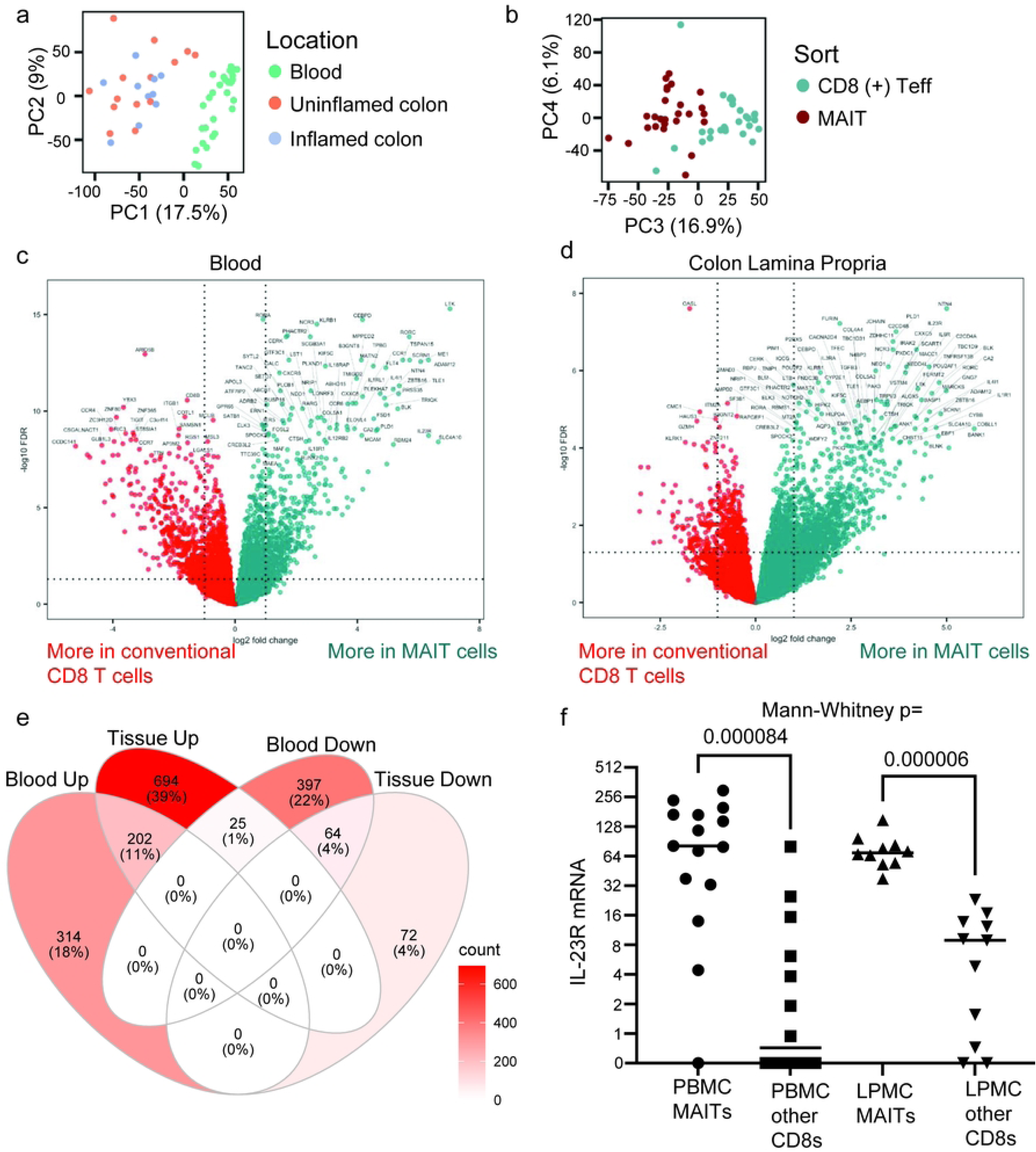
Blood and mucosal CD4^-^ MAIT cells have a transcriptome distinct from **other CD8 T cells.** Full transcriptome sequencing was performed on mRNA isolated from CD4^-^ MAIT or non-MAIT CD8^+^ T cells sorted from the blood or surgically resected colonic lamina propria of patients with or without IBD as an indication for surgery. Principal component (PC) analysis was performed on transcriptome profiles, separating anatomic source of cells in PCs 1 and 2 (**a**), and sorted cell phenotype in PCs 3 and 4 (**b**). Volcano plots of gene transcripts preferentially expressed in CD4^-^ MAIT cells (green) versus conventional CD8 T cells (red) are shown for cells sorted from blood (**c**) or colon (**d**). Dotted lines denote the threshold for significance, and the most differentially expressed genes are labeled by name. (**e**) Venn diagram showing the number of gene transcripts significantly more (“_Up”) or less (“_Down”) expressed in CD4^-^ MAIT cells relative to other CD8 T cells from PBMC (“Blood”) and/or colon (“Tissue”). (**f**) Normalized mRNA expression levels of the IL23R gene compared between CD4^-^ MAIT cells and non-MAIT CD8^+^ T cells isolated from blood (PBMC) or colon lamina propria (LPMC) by Mann-Whitney test.

Differential gene expression from normalized raw counts from all subjects revealed a number of genes up- or down-regulated in CD4^-^ MAIT cells relative to other CD8 cells in the blood (Fig 1**c**) or intestine (Fig 1**d**). A greater difference in gene expression was seen between CD4^-^ MAIT and other CD8 T cells in the blood than in the colon, both in terms of fold-change and significance, with the greatest difference observed in LTK, a receptor tyrosine kinase with no known ligand. Other genes up-regulated in CD4^-^ MAIT cells in this comparison encode several markers and factors known to be associated with both MAIT cells and Th17 cells, such as *RORC* (RORγt), *KLRB1* (CD161), *CCR6*, as well as the MAIT-associated gene *SLC4A10* (coding for NCBE, a sodium bicarbonate transporter). Components of the receptors for interleukins 18 (*IL18R1, IL18RAP*), 12 (*IL12RB2*) and 23 (*IL23R*) were also expressed more by CD4^-^ MAIT cells than other CD8 T cells. In contrast, CD4^-^ MAIT cells in the blood expressed comparatively less of the immunologically relevant *CD8B*, *CCR4*, *CCR7, VCAM1, KLRK1* (NKG2D), *GPR15, IFNG, CXCR3, GZMH* and *TIGIT* genes than other CD8 T cells.

In the colon, more genes were significantly up-then down-regulated in CD4^-^ MAIT cells relative to other CD8 T cells, with 202 of the up-regulated genes in the colon shared with those up-regulated by CD4^-^ MAIT cells in the blood (Fig 1**e**). The most significantly up-regulated colonic CD4^-^ MAIT gene was NTN4, encoding netrin 4, a laminin-related protein involved in neurite growth and migration, but with no known role in immunology. As in the blood, *IL23R* was among the more prominent genes differentially expressed in colonic CD4^-^ MAIT cells (Fig 1**f**), and other genes likewise associated with both MAIT cells and Th17 cells (*RORC*, *KLRB1* and *CCR6*) were again up-regulated, as in blood CD4^-^ MAIT cells. In contrast to blood, the cytokine receptor genes *IL18R1* and *IL12RB2* were not significantly more expressed by CD4^-^ MAIT cells in the colon, although this may simply reflect the smaller sample size of colonic MAIT cell transcriptomes.

Only 25 genes were up-regulated in CD4^-^ MAIT cells in the colon while being down-regulated in the blood, relative to other CD8 T cells. Of these, three have well-described immunological roles: *IL7* (the cytokine interleukin-7), *IL2RA* (CD25, the alpha chain of the receptor for interleukin-2), and *IFNGR2* (a non-ligand-binding beta chain of the gamma interferon receptor). Additionally, a more pronounced MAIT-specific expression of *IL1R1* (CD121a, the receptor for interleukin-1) was seen in the colon relative to blood, as was the gene encoding its signal transducing protein, *IRAK2*. Curiously, genes encoding some of the proteins involved in signal transduction by the B cell receptor (*BANK1, BLNK*) were also selectively up-regulated in CD4^-^ MAIT cells in the colon, while another (*BLK*) was MAIT-specific in both blood and colon.

Full transcriptome analysis is provided in Supplementary tables 2 and 3, and the normalized log_2_ counts in Supplementary table 4.

### Colonic MAIT cell immunophenotypes

Flow cytometry was performed on colonoscopic biopsies from 10 CD patients on no pharmaceutical treatment for CD (separately sampling both inflamed and uninflamed colonic segments from each) and 10 HC (screening colonoscopy recipients). Peripheral blood lymphocytes were also available for analyses from 16 of these subjects (8 per cohort).

We and others have previously reported that MAIT cells are less common in the blood of CD than HC subjects.[28, 71] In contrast, colonic CD3^+^, CD4^-^, CD161^+^, TCRVa7.2^+^ MAIT cells were a greater fraction of CD4^-^negative T cells in the colon in CD relative to HC, even if comparing only grossly uninflamed colon segments (*P* = 0.015, Fig 2**a**). Indeed, by paired analysis, there was no correlation between inflammation and this frequency of CD4^-^ MAIT cells in the colons of CD patients (*P* > 0.99). The presence of inflammation did, however, correlate with less NKG2D expression by CD4^-^ MAIT cells (Wilcoxon *P* = 0.004, Fig 2**b**), which also runs contrary to what has been described in blood.[28, 71] Regardless of inflammation and disease, CD4^-^ MAIT cells from the colon were predominantly CD8^+^ and CD103^+^ (Fig 2**c**, **d**). As the latter is the E-cadherin ligand which is mostly expressed in the epithelium, it suggests that most intestinal CD4^-^ MAIT cells, like many mucosal CD8^+^ T cells, are intra-epithelial lymphocytes (IEL). NKG2D and CD103 expression by conventional CD8^+^ T cells from these biopsies is shown for comparison (Fig 2**e**, **f**).

**Fig 2:**
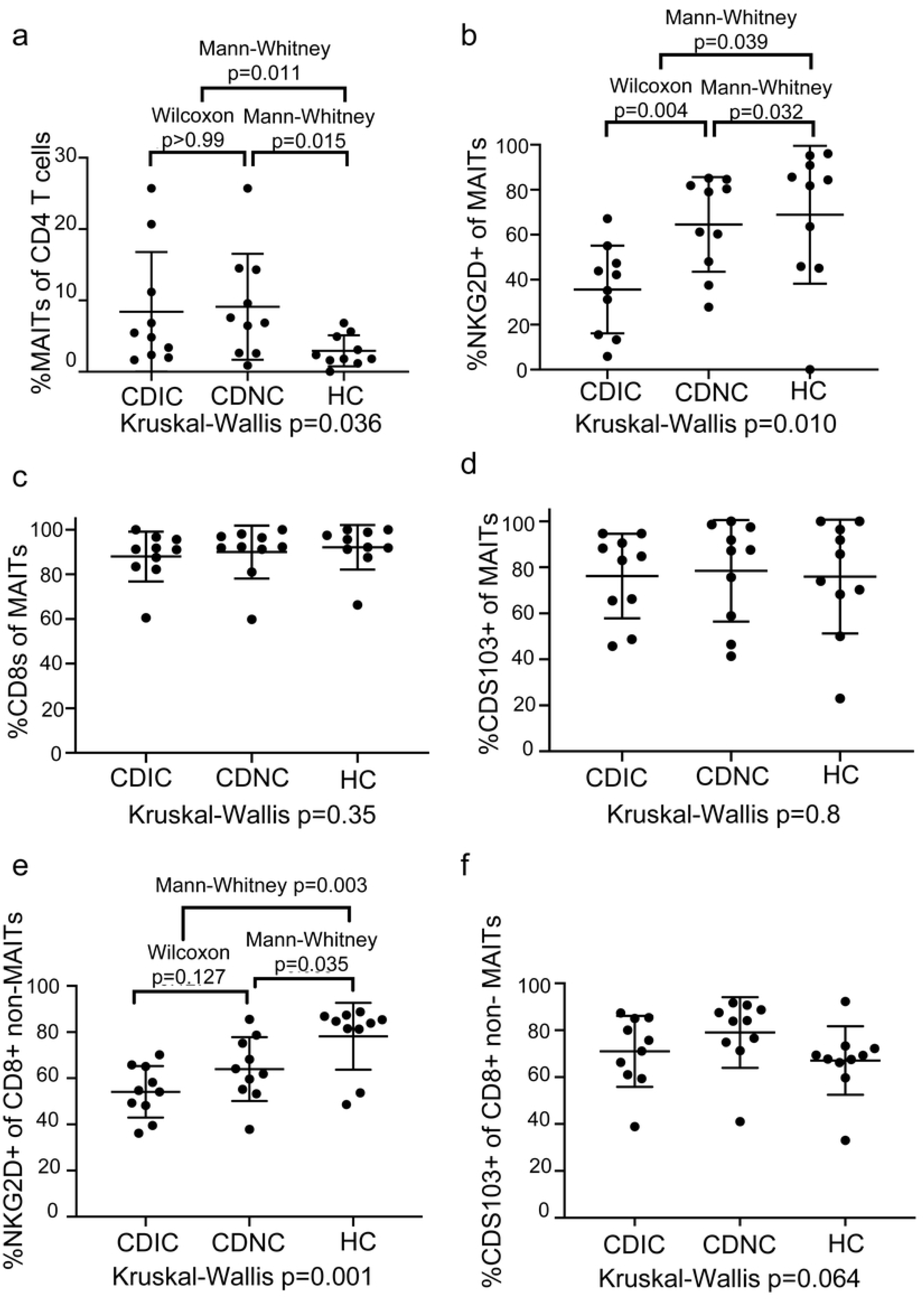
Frequency and phenotype of colonic CD4^-^ MAIT cells. (**a**) CD4^-^ MAIT cells from colon biopsies were quantified by flow cytometry as a percent of CD4^-^ negative T cells and found to be more common in biopsies from ten Crohn’s disease (CD) patients relative to ten healthy controls (HC), regardless of whether biopsied colon was inflamed (CDIC) or uninflamed (CDNC). The percent of colonic CD4^-^ MAIT cells expressing NKG2D (**b**), CD8 (**c**), or CD103 (**d**) and the percent of colonic CD8^+^ conventional T cells expressing NKG2D (**e**) or CD103 (**f**) on their surface was quantified by flow cytometry. A Kruskal-Wallis test for variance was performed for each parameter, and if it revealed a *P*-value less than 0.05, two-way paired (Wilcoxon) or unpaired (Mann-Whitney) non-parametric comparisons were performed as indicated between autologous or allogeneic specimens, respectively.

**Fig 3:**
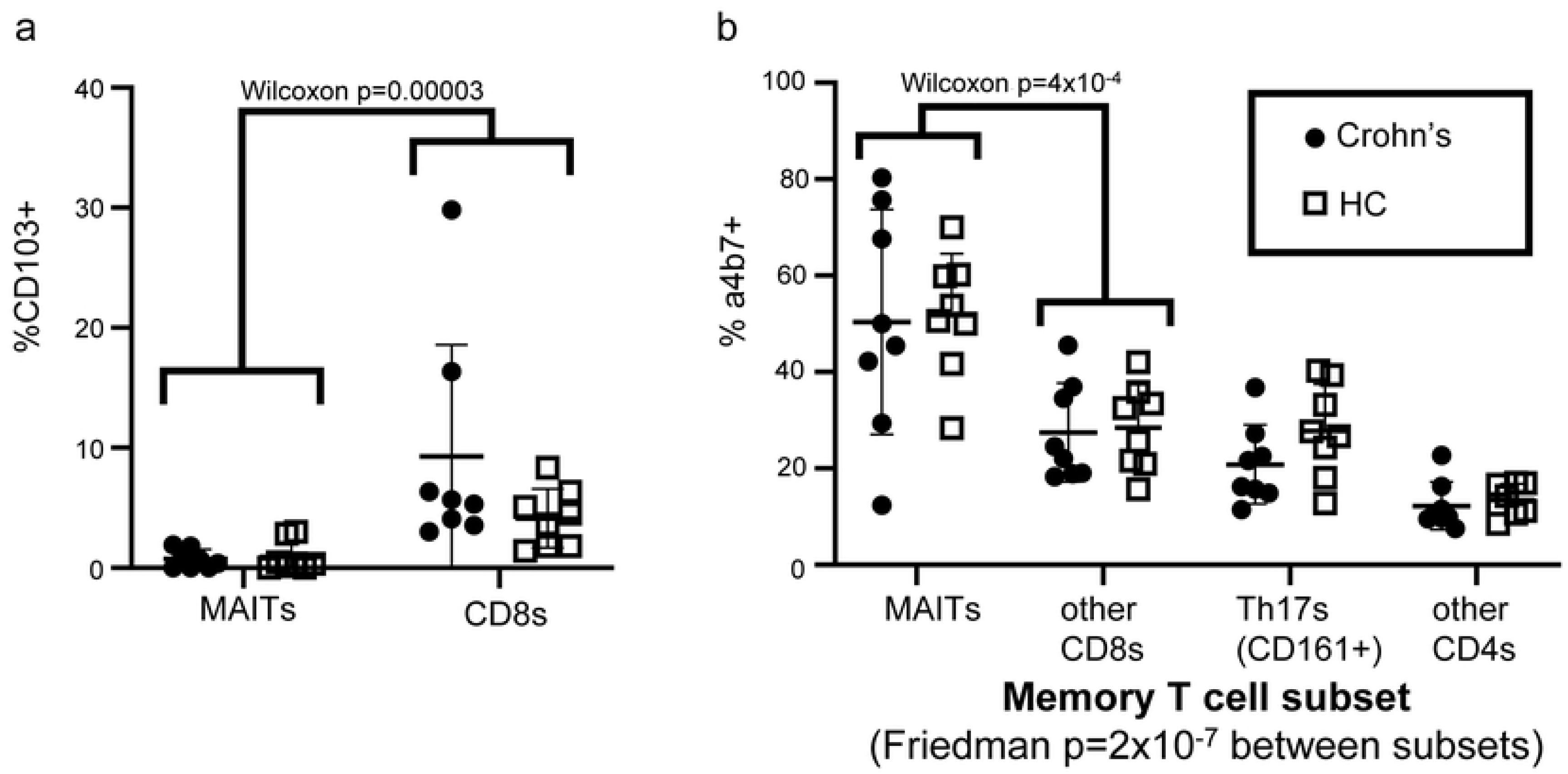
Frequency and phenotype of blood CD4^-^ MAIT cells. The percent of blood CD4^-^ MAIT or other T cell subsets (as indicated on x-axes) expressing CD103 (**a**) or integrin α4β7 (**b**) on their surface was quantified by flow cytometry. Pooling data from all cohorts, α4β7 expression showed highly significant variance between T cell subsets by paired analysis (Friedman test), with the most expression among CD4^-^ MAIT cells, which were significantly more α4β7^+^ (**b**) and less CD103^+^ (**a**) than other CD8 T cells by paired two-way (Wilcoxon) comparison. No differences in CD103 (**a**) or α4β7 expression (**b**) were seen between CD (black dots) and HC (open squares).

In contrast, CD103 (integrin αEβ7) expression by circulating CD4^-^ MAIT cells was rare, being significantly less common among circulating CD4^-^ MAIT cells than other antigen-experienced CD8^+^ T cells (Fig 3**a**). Conversely, CD4^-^ MAIT cells expressed significantly more of the gut-homing integrin α4β7 than any other antigen-experienced T cells, as previously described,[7] with no difference between CD and HC cohorts (Fig 3**b**).

### Colonic MAIT cell TCR sequences

To assess the clonality of MAIT cells in the colon, we FACS sorted each of up to 80 individual CD4^-^ MAIT cells per biopsy into its own well for paired TCR)alpha and beta chain sequencing from the above colonoscopic biopsies. Perhaps because CD161 expression is more ubiquitous in the colon than blood, flow cytometry alone did not produce a pure MAIT population, as less than half of the sorted cells had a canonical MAIT TCR alpha rearrangement of TRAV1-2/TRAJ12/20/33,[2-4, 72, 73] with TRAJ33 predominating in all (Supplementary table 5). Therefore, only productive paired TCR rearrangements of TRAV1-2 and TRAJ12/20/33 alpha chains were considered in further clonotype analysis. These alpha chains showed a consensus CDR3 amino acid sequence (CAV[M/R]DSNYQLIW) that was present in a majority of canonical MAIT cells (Fig 4**a**), while no clear consensus was evident in the CDR3 sequences of MAIT TCR beta chains. As described in the literature,[2-4, 72, 73] we observed a consistent selection of TCR beta variable genes in colonic CD4^-^ MAIT cells with TRBV6-4 or TRBV20-1 present in over half of CD4^-^ MAIT TCRs sequenced, as previously noted,[3] but regardless of disease or inflammation (Fig 4**b**). Additionally, in all cohorts we often found distinct cells with the same TCR alpha and beta chains (clonotypes) in a given biopsy, or common to both of two biopsies from different anatomic locations in the same person (i.e. both the inflamed and uninflamed biopsies from a CD patient) (Supplementary table 6 and Fig 5).

**Fig 4:**
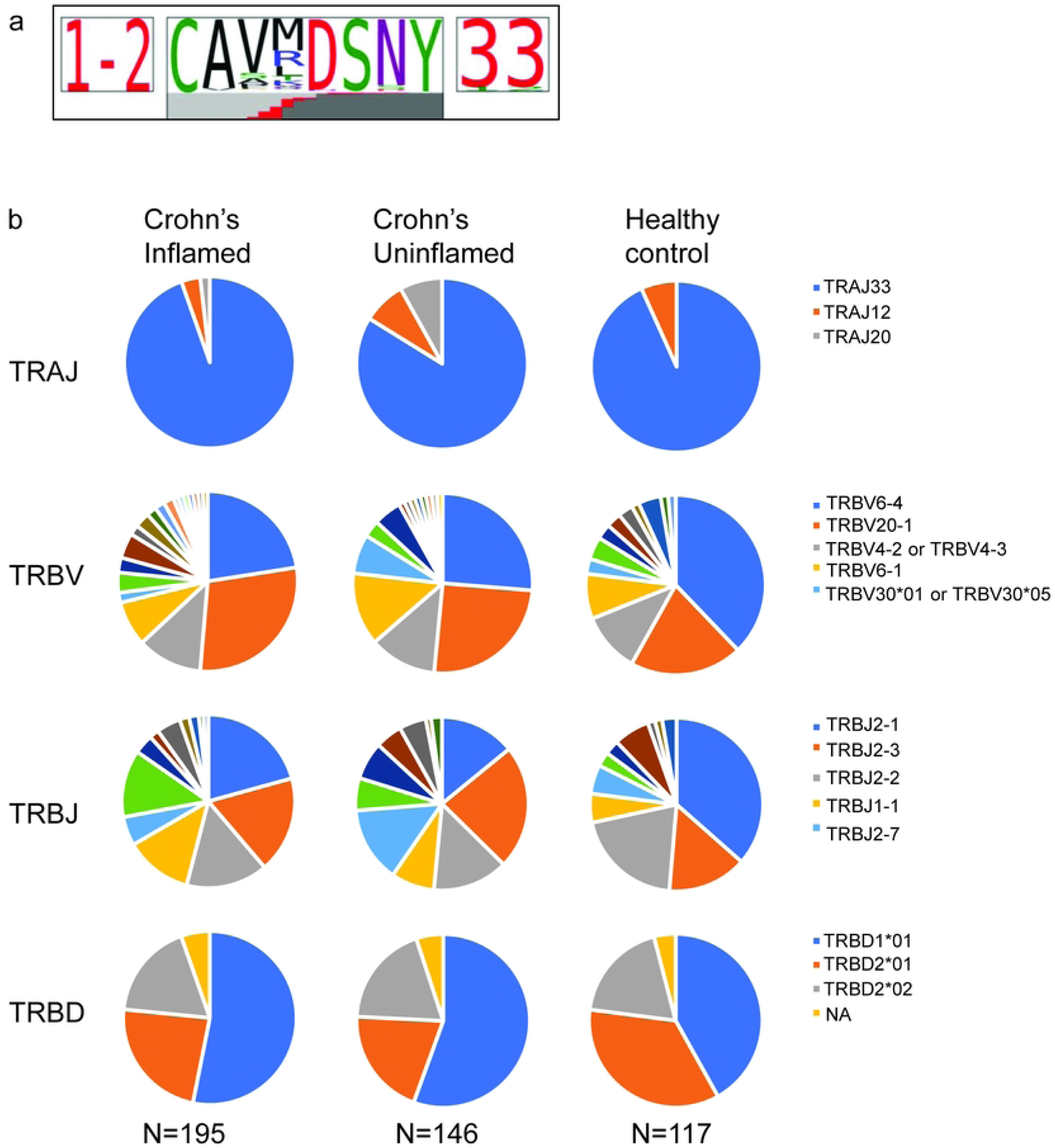
Colonic CD4^-^ MAIT cell TCR sequences. Full TCR alpha and beta chain gene transcripts were sequenced from individual CD4^-^ MAIT cells sorted from colon biopsies of the ten CD and ten HC subjects in Fig 2. Cells lacking the canonical MAIT TRAV1.2 sequence from their alpha chain were deemed contaminants and excluded from analyses. (**a**) Of all the remaining cells, a consensus sequence was observed in the TCR alpha chain CDR3 region. (**b**) The fraction of these CD4^-^ MAIT cells expressing different J-regions from the TCR alpha locus and different V, D, and J regions from the TCR beta locus is shown as a pie chart for inflamed or uninflamed colon biopsies from CD patients, or biopsies from healthy screening colonoscopy recipients.

**Fig 5:**
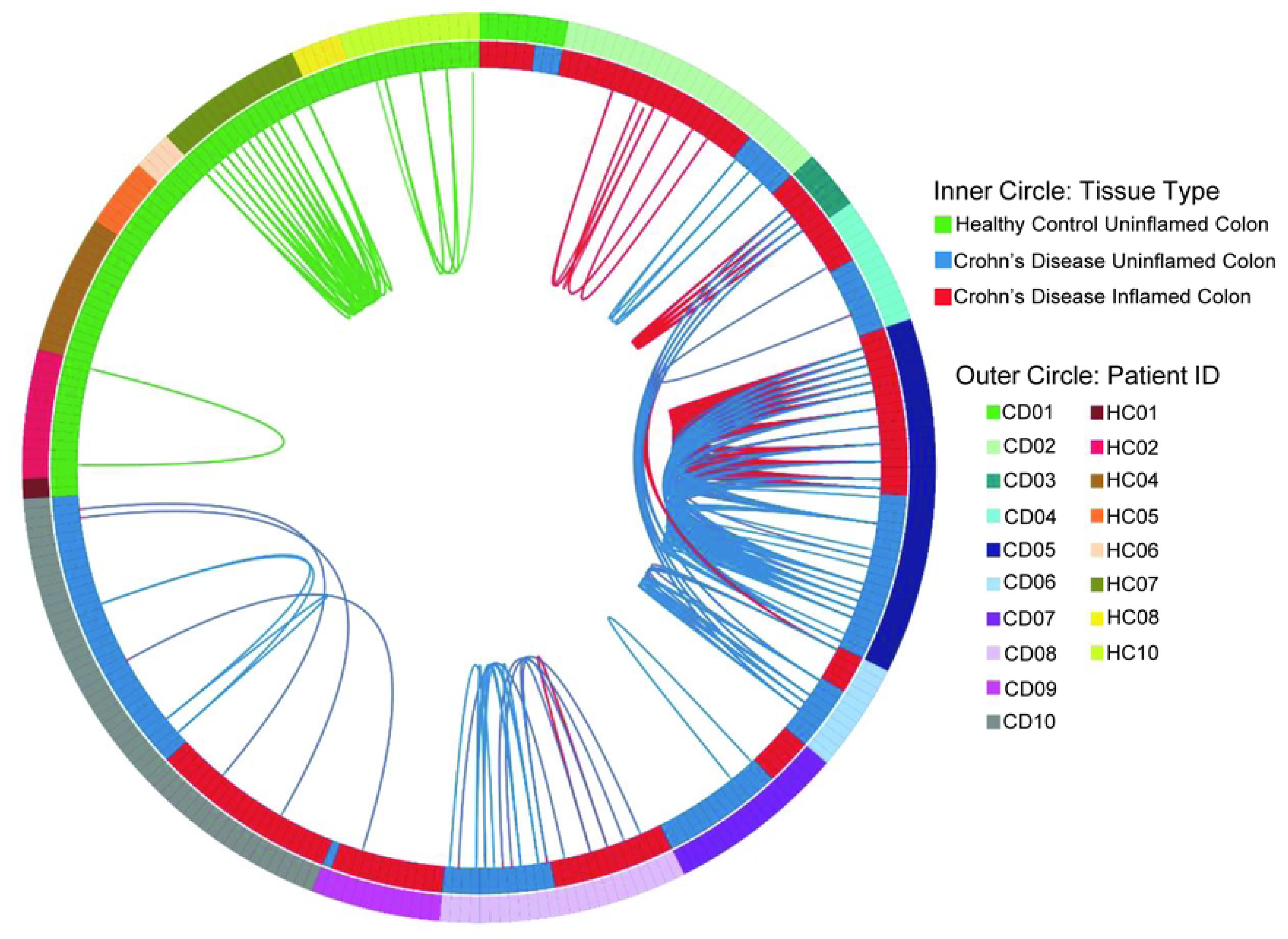
Colonic CD4^-^ MAIT cell clonality and overlap. Each individual CD4^-^ MAIT cell from Fig 3 with a canonical TRAV1.2 is shown as a point on the radius of this circos plot. Each individual from whom these MAIT cells were sorted is shown as a different color on the outer circle. The inner circle denotes whether MAIT cells were sorted from the colon biopsy of a HC (green) or a biopsy from the inflamed (red) or uninflamed (blue) colon of a CD patient. Any two MAIT cells with the exact same TCR alpha and beta sequences are connected by a thin line, colored to reflect their tissue of origin as in the inner circle (or purple, if cells from inflamed and uninflamed colon biopsies from CD patients have the same TCR).

Despite these TCRs being restricted to a finite range of molecular antigens presented by a genetically non-polymorphic molecule (MR1), there were few “public” TCRs seen in more than one person, none of which were from HC. Only two separate pairs of CD patients had any MAIT TCRs in common, indicating that convergent or pathogenic public clonotypes are not common in this disease.

CD4^-^ MAIT cells were also sorted from the PBMC of these patients, and total TCR beta chain CDR3 sequencing was performed on their DNA (Adaptive Biotechnologies). TCR diversity of circulating CD4^-^ MAIT cells in IBD was not significantly different in CD compared to HC (Fig 6**a**). The overlap in TCR repertoires between blood CD4^-^ MAIT cells was generally low, with little difference in overlap among or between CD and HC cohorts (Fig 6**b**, **c**). However, because up to 50,000 MAIT cells were analyzed per PBMC sample, we were able to identify 428 unique TCR beta chain CDR3 amino acid sequences that appeared in blood CD4^-^ MAIT cells from more than one person (Supplementary table 7), 103° of which were uniquely found among CD patients, and 39 of which were uniquely found in HC (the remaining 284 sequences being present in at least one person from each cohort).

**Fig 6:**
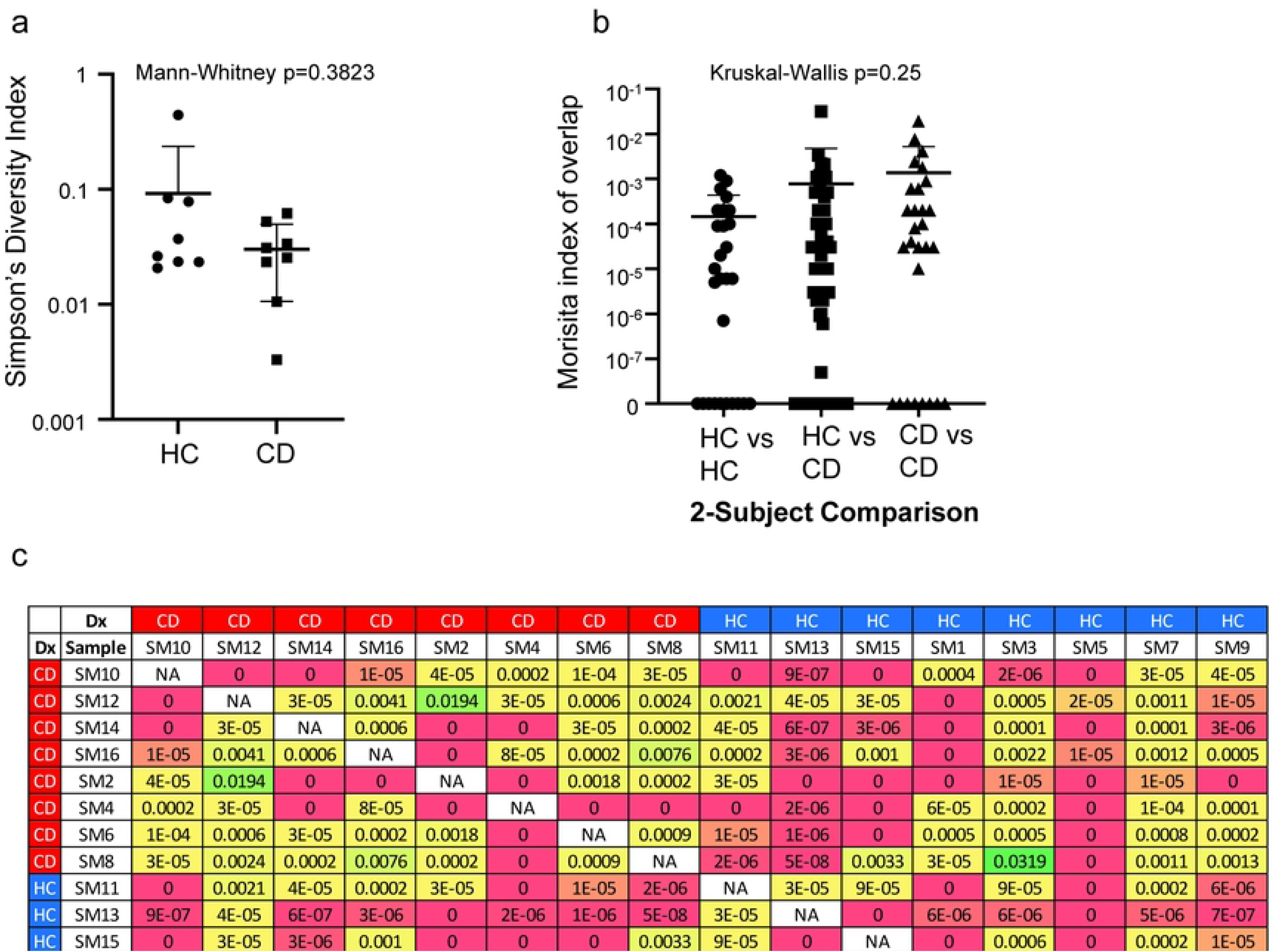
Blood CD4^-^ MAIT cell diversity and clonal overlap. CD4^-^ MAIT cells were sorted from the PBMC of eight Crohn’s disease (CD) patients or eight healthy controls (HC) at the time of colonoscopy from which they donated biopsies used in Figs 2-4, and their TCR beta chain CDR3 regions were sequenced. (**a**) The TCR beta repertoire diversity in each sample is plotted on a log scale, and the difference in Simpson’s diversity index (in which 1 denotes monoclonality and 0 means every cell has a different TCR beta sequence) between CD and HC samples shows no significance by unpaired non-parametric analysis (Mann-Whitney). (**b**) The Morisita index of overlap between TCR repertoires (in which 1 denotes complete overlap and 0 denotes no overlap) from each possible combination of any two of the 16 PBMC donors was calculated, and the overlap between every possible pairing is plotted on a log scale. The overlap between any two HC (HC vs HC), any two CD patients (CD vs CD) or any one HC and any one CD patient (HC vs CD) is shown, with unpaired non-parametric analysis (Kruskal-Wallis test) revealing no significant variance. (**c**) Actual Morisita indices of diversity plotted in (**b**) for each possible comparison between two CD (red border) and/or HC (blue border) subjects are shown and color coded on a grid.

Approximately half of the unique MAIT TCR beta sequences we found in biopsies were also found in CD4^-^ MAIT cells sorted from blood, with the vast majority of this overlap being between autologous samples from the same person. However, 13% of colonic CD4^-^ MAIT TCR beta sequences could also be found in the blood of another person, regardless of whether or not both the blood and colon biopsy donor had CD (Table 1). Thus, expanding our search of colonic CD4^-^ MAIT TCR beta sequences to also include blood revealed more colonic CD4^-^ MAIT TCR beta CDR3 sequences to be public (Supplementary table 8), but again did not find enrichment of these within a cohort to suggest pathogenic or protective clonotypes.

**Table 1:** Number of colonic MAIT cell TCRB sequences found, and their overlap with blood MAIT cells

| Biopsy | unique TCRβ CDR3 amino acid sequences | | | | % of total unique sequences | | |
| --- | --- | --- | --- | --- | --- | --- | --- |
|  | Total in colon | in both colon & blood | autologous overlap | allogeneic overlap | in both colon & blood | autologous overlap | allogeneic overlap |
| HC | 59 | 32 | 32 | 7 | 54% | 54% | 12% |
| CD all | 155 | 78 | 71 | 20 | 50% | 46% | 13% |
| CD NC | 78 | 47 | 41 | 16 | 60% | 53% | 21% |
| CD IC | 87 | 37 | 33 | 7 | 43% | 38% | 8% |
| total | 214 | 109 | 103 | 27 | 51% | 48% | 13% |

Abbreviations: TCR: T cell receptor, CDR: complementarity determining region, HC: healthy control, CD: Crohn's disease, NC: non-inflamed colon, IC: inflamed colon

## DISCUSSION

As their name would imply, mucosal-associated invariant T cells are part of the adaptive immune system at mucosal surfaces, with a function resembling the innate immune system. MAIT cells are known to respond to metabolite antigens produced by bacteria. Thus, as one of the largest mucosal surfaces in the human body, in contact with the highest density and diversity of bacteria in the human body, the intestinal mucosa is an organ whose MAIT cells warrant characterization. Given their similarity to Th17 cells, implicated in autoimmunity, MAIT cells may be of particular importance in spontaneous intestinal inflammation, such as CD. Indeed, two of the most effective treatment strategies for CD involve targeting integrin α4β7 and signaling through the IL-23 receptor, both of which we find highly enriched on CD4^-^ MAIT cells. We therefore compared gene expression in CD4^-^ MAIT cells to that of other CD8^+^ T cells in the intestines, as well as the blood, and compared blood and intestinal CD4^-^ MAIT surface protein immunophenotypes and TCR sequences in CD patients and HC.

Reciprocal to evidence that MAIT cells are a smaller fraction of T cells in the blood of CD patients, we corroborated evidence that these cells are enriched in their colons.[26–29] This suggests that MAIT cells are being sequestered from the blood to the intestinal mucosa in CD, as has been proposed to happen in the lungs with pulmonary tuberculosis.[8–10] We found an increased frequency of CD4^-^ MAIT cells in the intestinal mucosa of CD patients to be present even in uninflamed tissue, suggesting that MAIT cells predispose to inflammation, rather than being a consequence thereof. While this conflicts with an existing report finding MAIT cells to only be enriched in inflamed colon,[29] that study pooled data from CD and UC patients, and also was unable to examine biopsies with more than mild inflammation, which could have overlapped with specimens deemed endoscopically uninflamed in our study. Furthermore, it did not exclude CD4^+^ T cells, from which a high frequency of CD161^+^ Th17 cells present in the intestinal mucosa[69]could contaminate MAIT cells. In contrast, the fraction of intestinal CD4^-^ MAIT cells expressing NKG2D was lower in inflamed than uninflamed colon, suggesting that inflammation leads to its down-regulation. This is consistent with IL-15-driven NKG2D expression by memory CD8 T (including MAIT) cells being inhibited by TCR activation.[74] While it is possible that a relative lack NKG2D on MAIT cells could be a cause rather than an effect of local intestinal inflammation, this hypothesis would run contrary to the observation that NKG2D blockade reduced CD clinical activity by week 12 in a randomized trial.[75]

We also found that the vast majority of mucosal CD4^-^ MAIT cells express CD103 (integrin αEβ7), which adheres to the E-cadherin molecule expressed by intestinal epithelial cells.[76] Although analyses of histological morphology were outside the scope of this study, the CD103^+^ phenotype of intestinal MAIT cells suggests that they are intra-epithelial lymphocytes (IEL), in close proximity to the intestinal lumen, and thus the gut microbiome. A thick layer of mucous produced by the intestinal mucosa limits exposure of the gut epithelium to much of the microbiome itself. However, the riboflavin precursors uniquely recognized by MAIT cells are small organic molecules which may more readily diffuse across mucous than an intact microorganism or its protein antigens. Thus, as IEL, MAIT cells would be uniquely suited to monitor the microbiome through its metabolic products.

Although human MAIT cell clonal diversity has been studied previously in the blood,[3, 72, 73, 77] little is known about it at the human colonic mucosal barrier, where TCR diversity is much narrower.[69] We therefore had hypothesized that intestinal MAIT cells would be pauciclonal. Furthermore, because they are restricted by the genetically non-polymorphic MR1 molecule,[7] and recognize a finite range of riboflavin precursors.[6] we hypothesized that such pauciclonality would include a large number of “public” TCR sequences common to multiple individuals. If so, the presence or absence of specific TCR sequences in the colons of CD patients relative to HC would suggest that specific MR1-restricted antigens are either pathogenic or protective, respectively, in IBD. While the antigens recognized by the colonic CD4^-^ MAIT cells studied in this report are unknown, the consensus amino acid motif of the TCR alpha chain CDR3s we report (CAV[M/R]DSNYQLIW) most resembles a sequence (CAVRDS[N/D]Y[K/Q]L[S/I]) reported for peripheral blood MAIT cells that respond *in vitro* to *Candida albicans* antigens.[77] Although not known to be related to disease pathogenesis, *C. albicans* is an immunogen for the anti-Saccharomyces cerevisiae antibody (ASCA)[78] that has been sufficiently associated with CD to serve as a clinical marker of disease since the 1990’s[79] and may predict a more aggressive disease course.[80] Thus, further comparison of MAIT TCR sequences between CD patients with and without high ASCA titers may reveal an immunophenotypic difference between these IBD patient cohorts.

After restricting analyses to T cells containing the canonical alpha chains with which MAIT cells are defined, we found the corresponding beta chains to be surprisingly diverse, with more than half of all colonic CD4^-^ MAIT cells analyzed containing a unique TCR beta chain CDR3 hypervariable sequence. Of those colonic MAIT sequences found in more than one cell, only a small minority were shared between different individuals (i.e. “public”), and never among the HC subjects we analyzed. Those few “public” TCR beta sequences we identified between colon biopsies from different CD patients could suggest potentially pathogenic antigen specificities, but none of them were shared between more than two patients, so any hypothesized pathogenic specificity would not be a widespread characteristic of this disease. In individual CD patients, the same TCR beta sequence could often be found in MAIT cells that were harvested from both the inflamed and uninflamed colon, suggesting that their antigen specificity is not correlated with tissue inflammation.

“Public” TCR beta chain CDR3 sequences were also uncommon in the peripheral blood, but more readily found due to the much larger number of MAIT cells that could be sorted from blood than biopsies. More than twice as many unique public sequences were exclusive to CD patients as were exclusive to HC. However, more than half of all public sequences were seen in both CD and HC cohorts, suggesting that the observed exclusivity was more a consequence of random chance than any proof that pathogenic (i.e. CD-exclusive) or protective (i.e. HC-exclusive) MAIT clonotypes exist. Indeed, many of the unique colonic CD4^-^ MAIT TCR beta chain CDR3 sequences seen exclusively in CD patients could also be found in the peripheral blood of HC, and vice versa. Given that the diversity of CD4^-^ MAIT TCR beta chain sequences appears to greatly exceed the diversity of small, organic molecules in the riboflavin synthesis pathway that can be recognized by MAIT cells, categorization of these beta chain sequences by their ability to bind specific MR1-presented antigens may better clarify similarities and differences between CD and HC. Another possibility that explains the vast TCR β-chain repertoire is that there is little requirement for a particular β-chain to recognize the same antigen with most of the recognition being provided by the TCR α-chain.

In summary, we evaluated CD4^-^ MAIT cells from the blood and colon of people with and without CD, by flow cytometry, mRNA expression, and TCR sequencing. We found CD4^-^ MAIT cells to be concentrated in the colon in CD relative to HC, regardless of whether or not they came from an inflamed colon segment. Indeed, we found that CD4^-^ MAIT cells in the blood express more of the gut-homing integrin α4β7 than other CD8 T cells, indicating a tropism for the intestinal mucosa. As integrin α4β7 has become a major therapeutic target in the treatment of IBD, it remains to be seen how a response to anti-integrin therapy affects, or may even be predicted by, MAIT cells. Likewise, we report that colonic CD4^-^ MAIT cells express much more of the IL-23 receptor than do other CD8 T cells. As IL-23 blockade is also now a major treatment strategy for IBD, a study of MAIT cells before and during anti-IL-23 therapy could also identify biomarkers with which to optimize treatment or predict its efficacy. While colonic CD4^-^

MAIT cells proved far more polymorphic at the TCR/clonal level than anticipated, with few public TCR sequences to correlate with health or disease, we did observe considerable MAIT TCR repertoire overlap between different anatomic locations within an individual, suggesting an individual would likely also show overlap in their MAIT repertoire at different time points.

Longitudinally tracking MAIT TCR repertoire over time in an individual, such as before and during anti-integrin or anti-IL-23 therapy, would provide further insight into how IBD therapeutics work, correlate MAIT cell specificity with disease course, and hence reveal novel insights into the pathogenesis of IBD.

## Data Availability

All relevant data are within the manuscript and its Supporting Information files.

## Acknowledgements

We thank Kassidy Benoscek for subject recruitment, Adam Wojno for assistance with flow cytometry, Hannah DeBerg for assistance with R programming, and Brenda Norris, Virginia Greene, and Taylor Lawson for assistance with manuscript preparation.

## REFERENCES

1. Billerbeck E, Kang YH, Walker L, Lockstone H, Grafmueller S, Fleming V, et al. Analysis of CD161 expression on human CD8+ T cells defines a distinct functional subset with tissue-homing properties. Proc Natl Acad Sci U S A. 2010;107(7):3006–11. Epub 2010/02/06. doi: 10.1073/pnas.0914839107. PubMed PMID: 20133607; PubMed Central PMCID: PMCPMC2840308.

2. Gherardin NA, Souter MN, Koay HF, Mangas KM, Seemann T, Stinear TP, et al. Human blood MAIT cell subsets defined using MR1 tetramers. Immunol Cell Biol. 2018;96(5):507–25. Epub 2018/02/14. doi: 10.1111/imcb.12021. PubMed PMID: 29437263; PubMed Central PMCID: PMCPMC6446826.

3. Tilloy F, Treiner E, Park SH, Garcia C, Lemonnier F, de la Salle H, et al. An invariant T cell receptor alpha chain defines a novel TAP-independent major histocompatibility complex class Ib-restricted alpha/beta T cell subpopulation in mammals. J Exp Med. 1999;189(12):1907–21.

4. Porcelli S, Yockey CE, Brenner MB, Balk SP. Analysis of T cell antigen receptor (TCR) expression by human peripheral blood CD4-8-alpha/beta T cells demonstrates preferential use of several V beta genes and an invariant TCR alpha chain. J Exp Med. 1993;178(1):1–16. doi: 10.1084/jem.178.1.1. PubMed PMID: 8391057; PubMed Central PMCID: PMCPMC2191070.

5. D’Souza C, Pediongco T, Wang H, Scheerlinck JY, Kostenko L, Esterbauer R, et al. Mucosal-Associated Invariant T Cells Augment Immunopathology and Gastritis in Chronic Helicobacter pylori Infection. J Immunol. 2018;200(5):1901–16. Epub 2018/01/31. doi: 10.4049/jimmunol.1701512. PubMed PMID: 29378910.

6. Kjer-Nielsen L, Patel O, Corbett AJ, Le NJ, Meehan B, Liu L, et al. MR1 presents microbial vitamin B metabolites to MAIT cells. Nature. 2012;491(7426):717–23. doi: nature11605 [pii];10.1038/nature11605 [doi].

7. Treiner E, Duban L, Bahram S, Radosavljevic M, Wanner V, Tilloy F, et al. Selection of evolutionarily conserved mucosal-associated invariant T cells by MR1. Nature. 2003;422(6928):164–9. doi: 10.1038/nature01433 [doi];nature01433 [pii].

8. Dusseaux M, Martin E, Serriari N, Peguillet I, Premel V, Louis D, et al. Human MAIT cells are xenobiotic-resistant, tissue-targeted, CD161hi IL-17-secreting T cells. Blood. 2011;117(4):1250–9. doi: blood-2010-08-303339 [pii];10.1182/blood-2010-08-303339 [doi].

9. Coulter F, Parrish A, Manning D, Kampmann B, Mendy J, Garand M, et al. IL-17 Production from T Helper 17, Mucosal-Associated Invariant T, and gammadelta Cells in Tuberculosis Infection and Disease. Front Immunol. 2017;8:1252. Epub 2017/10/28. doi: 10.3389/fimmu.2017.01252. PubMed PMID: 29075255; PubMed Central PMCID: PMCPMC5641565.

10. Gold MC, Cerri S, Smyk-Pearson S, Cansler ME, Vogt TM, Delepine J, et al. Human mucosal associated invariant T cells detect bacterially infected cells. PLoS Biol. 2010;8(6):e1000407. doi: 10.1371/journal.pbio.1000407 [doi].

11. Kennedy-Nasser AA, Ku S, Castillo-Caro P, Hazrat Y, Wu MF, Liu H, et al. Ultra low-dose IL-2 for GVHD prophylaxis after allogeneic hematopoietic stem cell transplantation mediates expansion of regulatory T cells without diminishing antiviral and antileukemic activity. Clin Cancer Res. 2014;20(8):2215–25. doi: 1078-0432.CCR-13-3205 [pii];10.1158/1078-0432.CCR-13-3205 [doi].

12. Salerno-Goncalves R, Luo D, Fresnay S, Magder L, Darton TC, Jones C, et al. Challenge of Humans with Wild-type Salmonella enterica Serovar Typhi Elicits Changes in the Activation and Homing Characteristics of Mucosal-Associated Invariant T Cells. Front Immunol. 2017;8:398. Epub 2017/04/22. doi: 10.3389/fimmu.2017.00398. PubMed PMID: 28428786; PubMed Central PMCID: PMCPMC5382150.

13. Leeansyah E, Ganesh A, Quigley MF, Sonnerborg A, Andersson J, Hunt PW, et al. Activation, exhaustion, and persistent decline of the antimicrobial MR1-restricted MAIT-cell population in chronic HIV-1 infection. Blood. 2013;121(7):1124–35. Epub 2012/12/18. doi: 10.1182/blood-2012-07-445429. PubMed PMID: 23243281; PubMed Central PMCID: PMCPMC3575756.

14. Khaitan A, Kilberg M, Kravietz A, Ilmet T, Tastan C, Mwamzuka M, et al. HIV-Infected Children Have Lower Frequencies of CD8+ Mucosal-Associated Invariant T (MAIT) Cells that Correlate with Innate, Th17 and Th22 Cell Subsets. PLoS One. 2016;11(8):e0161786. Epub 2016/08/26. doi: 10.1371/journal.pone.0161786. PubMed PMID: 27560150; PubMed Central PMCID: PMCPMC4999196.

15. Hengst J, Strunz B, Deterding K, Ljunggren HG, Leeansyah E, Manns MP, et al. Nonreversible MAIT cell-dysfunction in chronic hepatitis C virus infection despite successful interferon-free therapy. Eur J Immunol. 2016;46(9):2204–10. Epub 2016/06/15. doi: 10.1002/eji.201646447. PubMed PMID: 27296288.

16. Bolte FJ, O’Keefe AC, Webb LM, Serti E, Rivera E, Liang TJ, et al. Intra-Hepatic Depletion of Mucosal-Associated Invariant T Cells in Hepatitis C Virus-Induced Liver Inflammation. Gastroenterology. 2017;153(5):1392–403 e2. Epub 2017/08/07. doi: 10.1053/j.gastro.2017.07.043. PubMed PMID: 28780074; PubMed Central PMCID: PMCPMC5669813.

17. Grimaldi D, Le Bourhis L, Sauneuf B, Dechartres A, Rousseau C, Ouaaz F, et al. Specific MAIT cell behaviour among innate-like T lymphocytes in critically ill patients with severe infections. Intensive Care Med. 2014;40(2):192–201. Epub 2013/12/11. doi: 10.1007/s00134-013-3163-x. PubMed PMID: 24322275.

18. Hannaway RF, Wang X, Schneider M, Slow S, Cowan J, Brockway B, et al. Mucosal-associated invariant T cells and Vdelta2(+) gammadelta T cells in community acquired pneumonia: association of abundance in sputum with clinical severity and outcome. Clin Exp Immunol. 2020;199(2):201–15. Epub 2019/10/07. doi: 10.1111/cei.13377. PubMed PMID: 31587268; PubMed Central PMCID: PMCPMC6954682.

19. Miyazaki Y, Miyake S, Chiba A, Lantz O, Yamamura T. Mucosal-associated invariant T cells regulate Th1 response in multiple sclerosis. Int Immunol. 2011;23(9):529–35. doi: dxr047 [pii];10.1093/intimm/dxr047 [doi].

20. Berglin L, Bergquist A, Johansson H, Glaumann H, Jorns C, Lunemann S, et al. In situ characterization of intrahepatic non-parenchymal cells in PSC reveals phenotypic patterns associated with disease severity. PLoS One. 2014;9(8):e105375. doi: 10.1371/journal.pone.0105375 [doi];PONE-D-14-20860 [pii].

21. Wang JJ, Macardle C, Weedon H, Beroukas D, Banovic T. Mucosal-associated invariant T cells are reduced and functionally immature in the peripheral blood of primary Sjogren’s syndrome patients. Eur J Immunol. 2016;46(10):2444–53. Epub 2016/07/28. doi: 10.1002/eji.201646300. PubMed PMID: 27461134.

22. Witte E, Witte K, Warszawska K, Sabat R, Wolk K. Interleukin-22: a cytokine produced by T, NK and NKT cell subsets, with importance in the innate immune defense and tissue protection. Cytokine Growth Factor Rev. 2010;21(5):365–79. Epub 2010/09/28. doi: 10.1016/j.cytogfr.2010.08.002. PubMed PMID: 20870448.

23. Magalhaes I, Pingris K, Poitou C, Bessoles S, Venteclef N, Kiaf B, et al. Mucosal-associated invariant T cell alterations in obese and type 2 diabetic patients. J Clin Invest. 2015;125(4):1752–62. Epub 2015/03/10. doi: 10.1172/JCI78941. PubMed PMID: 25751065; PubMed Central PMCID: PMCPMC4396481.

24. Carolan E, Tobin LM, Mangan BA, Corrigan M, Gaoatswe G, Byrne G, et al. Altered distribution and increased IL-17 production by mucosal-associated invariant T cells in adult and childhood obesity. J Immunol. 2015;194(12):5775–80. Epub 2015/05/17. doi: 10.4049/jimmunol.1402945. PubMed PMID: 25980010.

25. Dunne MR, Elliott L, Hussey S, Mahmud N, Kelly J, Doherty DG, et al. Persistent changes in circulating and intestinal gammadelta T cell subsets, invariant natural killer T cells and mucosal-associated invariant T cells in children and adults with coeliac disease. PLoS One. 2013;8(10):e76008. Epub 2013/10/15. doi: 10.1371/journal.pone.0076008. PubMed PMID: 24124528; PubMed Central PMCID: PMCPMC3790827.

26. Hiejima E, Kawai T, Nakase H, Tsuruyama T, Morimoto T, Yasumi T, et al. Reduced Numbers and Proapoptotic Features of Mucosal-associated Invariant T Cells as a Characteristic Finding in Patients with Inflammatory Bowel Disease. Inflamm Bowel Dis. 2015;21(7):1529–40. Epub 2015/05/07. doi: 10.1097/MIB.0000000000000397. PubMed PMID: 25946569.

27. Haga K, Chiba A, Shibuya T, Osada T, Ishikawa D, Kodani T, et al. MAIT cells are activated and accumulated in the inflamed mucosa of ulcerative colitis. J Gastroenterol Hepatol. 2016;31(5):965–72. Epub 2015/11/22. doi: 10.1111/jgh.13242. PubMed PMID: 26590105.

28. Serriari NE, Eoche M, Lamotte L, Lion J, Fumery M, Marcelo P, et al. Innate mucosal-associated invariant T (MAIT) cells are activated in inflammatory bowel diseases. Clin Exp Immunol. 2014;176(2):266–74. doi: 10.1111/cei.12277 [doi].

29. Tominaga K, Yamagiwa S, Setsu T, Kimura N, Honda H, Kamimura H, et al. Possible involvement of mucosal-associated invariant T cells in the progression of inflammatory bowel diseases. Biomed Res. 2017;38(2):111–21. doi: 10.2220/biomedres.38.111. PubMed PMID: 28442662.

30. Willing A, Leach OA, Ufer F, Attfield KE, Steinbach K, Kursawe N, et al. CD8(+) MAIT cells infiltrate into the CNS and alterations in their blood frequencies correlate with IL-18 serum levels in multiple sclerosis. Eur J Immunol. 2014;44(10):3119–28. Epub 2014/07/22. doi: 10.1002/eji.201344160. PubMed PMID: 25043505.

31. Salou M, Nicol B, Garcia A, Baron D, Michel L, Elong-Ngono A, et al. Neuropathologic, phenotypic and functional analyses of Mucosal Associated Invariant T cells in Multiple Sclerosis. Clin Immunol. 2016;166–167:1–11. Epub 2016/04/07. doi: 10.1016/j.clim.2016.03.014. PubMed PMID: 27050759.

32. Annibali V, Ristori G, Angelini DF, Serafini B, Mechelli R, Cannoni S, et al. CD161(high)CD8+T cells bear pathogenetic potential in multiple sclerosis. Brain. 2011;134(Pt 2):542–54. Epub 2011/01/11. doi: 10.1093/brain/awq354. PubMed PMID: 21216829.

33. Abrahamsson SV, Angelini DF, Dubinsky AN, Morel E, Oh U, Jones JL, et al. Non-myeloablative autologous haematopoietic stem cell transplantation expands regulatory cells and depletes IL-17 producing mucosal-associated invariant T cells in multiple sclerosis. Brain. 2013;136(Pt 9):2888–903. Epub 2013/07/19. doi: 10.1093/brain/awt182. PubMed PMID: 23864273; PubMed Central PMCID: PMCPMC3754461.

34. Price AB, Morson BC. Inflammatory bowel disease: the surgical pathology of Crohn’s disease and ulcerative colitis. Hum Pathol. 1975;6(1):7–29. Epub 1975/01/01. doi: 10.1016/s0046-8177(75)80107-9. PubMed PMID: 1089084.

35. Ehlers S. Why does tumor necrosis factor targeted therapy reactivate tuberculosis? J Rheumatol Suppl. 2005;74:35–9. Epub 2005/03/03. PubMed PMID: 15742463.

36. Agrawal G, Aitken J, Hamblin H, Collins M, Borody TJ. Putting Crohn’s on the MAP: Five Common Questions on the Contribution of Mycobacterium avium subspecies paratuberculosis to the Pathophysiology of Crohn’s Disease. Dig Dis Sci. 2021;66(2):348–58. Epub 2020/10/23. doi: 10.1007/s10620-020-06653-0. PubMed PMID: 33089484; PubMed Central PMCID: PMCPMC7577843.

37. Jostins L, Ripke S, Weersma RK, Duerr RH, McGovern DP, Hui KY, et al. Host-microbe interactions have shaped the genetic architecture of inflammatory bowel disease. Nature. 2012;491(7422):119–24. doi: nature11582 [pii];10.1038/nature11582 [doi].

38. Brand S. Crohn’s disease: Th1, Th17 or both? The change of a paradigm: new immunological and genetic insights implicate Th17 cells in the pathogenesis of Crohn’s disease. Gut. 2009;58(8):1152–67. Epub 2009/07/14. doi: 10.1136/gut.2008.163667. PubMed PMID: 19592695.

39. Galvez J. Role of Th17 Cells in the Pathogenesis of Human IBD. ISRN Inflamm. 2014;2014:928461. Epub 2014/08/08. doi: 10.1155/2014/928461. PubMed PMID: 25101191; PubMed Central PMCID: PMCPMC4005031.

40. Yasuda K, Takeuchi Y, Hirota K. The pathogenicity of Th17 cells in autoimmune diseases. Semin Immunopathol. 2019;41(3):283–97. Epub 2019/03/21. doi: 10.1007/s00281-019-00733-8. PubMed PMID: 30891627.

41. Han L, Yang J, Wang X, Li D, Lv L, Li B. Th17 cells in autoimmune diseases. Front Med. 2015;9(1):10–9. Epub 2015/02/06. doi: 10.1007/s11684-015-0388-9. PubMed PMID: 25652649.

42. Maddur MS, Miossec P, Kaveri SV, Bayry J. Th17 cells: biology, pathogenesis of autoimmune and inflammatory diseases, and therapeutic strategies. Am J Pathol. 2012;181(1):8–18. doi: S0002-9440(12)00336-7 [pii];10.1016/j.ajpath.2012.03.044 [doi].

43. Huang S, Martin E, Kim S, Yu L, Soudais C, Fremont DH, et al. MR1 antigen presentation to mucosal-associated invariant T cells was highly conserved in evolution. Proc Natl Acad Sci U S A. 2009;106(20):8290–5. Epub 2009/05/07. doi: 10.1073/pnas.0903196106. PubMed PMID: 19416870; PubMed Central PMCID: PMCPMC2688861.

44. Koay HF, Gherardin NA, Enders A, Loh L, Mackay LK, Almeida CF, et al. A three-stage intrathymic development pathway for the mucosal-associated invariant T cell lineage. Nat Immunol. 2016;17(11):1300–11. Epub 2016/10/21. doi: 10.1038/ni.3565. PubMed PMID: 27668799.

45. Bengsch B, Seigel B, Flecken T, Wolanski J, Blum HE, Thimme R. Human Th17 cells express high levels of enzymatically active dipeptidylpeptidase IV (CD26). J Immunol. 2012;188(11):5438–47. Epub 2012/04/28. doi: 10.4049/jimmunol.1103801. PubMed PMID: 22539793.

46. Brozova J, Karlova I, Novak J. Analysis of the Phenotype and Function of the Subpopulations of Mucosal-Associated Invariant T Cells. Scand J Immunol. 2016;84(4):245–51. Epub 2016/07/31. doi: 10.1111/sji.12467. PubMed PMID: 27474379.

47. Mpina M, Maurice NJ, Yajima M, Slichter CK, Miller HW, Dutta M, et al. Controlled Human Malaria Infection Leads to Long-Lasting Changes in Innate and Innate-like Lymphocyte Populations. J Immunol. 2017;199(1):107–18. Epub 2017/06/04. doi: 10.4049/jimmunol.1601989. PubMed PMID: 28576979; PubMed Central PMCID: PMCPMC5528886.

48. Leeansyah E, Loh L, Nixon DF, Sandberg JK. Acquisition of innate-like microbial reactivity in mucosal tissues during human fetal MAIT-cell development. Nat Commun. 2014;5:3143. Epub 2014/01/24. doi: 10.1038/ncomms4143. PubMed PMID: 24452018; PubMed Central PMCID: PMCPMC3916833.

49. Brahmer JR, Drake CG, Wollner I, Powderly JD, Picus J, Sharfman WH, et al. Phase I study of single-agent anti-programmed death-1 (MDX-1106) in refractory solid tumors: safety, clinical activity, pharmacodynamics, and immunologic correlates. J Clin Oncol. 2010;28(19):3167–75. doi: JCO.2009.26.7609 [pii];10.1200/JCO.2009.26.7609 [doi].

50. Leeansyah E, Svard J, Dias J, Buggert M, Nystrom J, Quigley MF, et al. Arming of MAIT Cell Cytolytic Antimicrobial Activity Is Induced by IL-7 and Defective in HIV-1 Infection. PLoS Pathog. 2015;11(8):e1005072. Epub 2015/08/22. doi: 10.1371/journal.ppat.1005072. PubMed PMID: 26295709; PubMed Central PMCID: PMCPMC4546682.

51. Tang XZ, Jo J, Tan AT, Sandalova E, Chia A, Tan KC, et al. IL-7 licenses activation of human liver intrasinusoidal mucosal-associated invariant T cells. J Immunol. 2013;190(7):3142-doi: jimmunol.1203218 [pii];10.4049/jimmunol.1203218 [doi].

52. Ussher JE, Bilton M, Attwod E, Shadwell J, Richardson R, de Lara C, et al. CD161++ CD8+ T cells, including the MAIT cell subset, are specifically activated by IL-12+IL-18 in a TCR-independent manner. Eur J Immunol. 2014;44(1):195–203. Epub 2013/09/11. doi: 10.1002/eji.201343509. PubMed PMID: 24019201; PubMed Central PMCID: PMCPMC3947164.

53. Sattler A, Dang-Heine C, Reinke P, Babel N. IL-15 dependent induction of IL-18 secretion as a feedback mechanism controlling human MAIT-cell effector functions. Eur J Immunol. 2015;45(8):2286–98. Epub 2015/06/06. doi: 10.1002/eji.201445313. PubMed PMID: 26046663.

54. Zhou L, Ivanov, II, Spolski R, Min R, Shenderov K, Egawa T, et al. IL-6 programs T(H)-17 cell differentiation by promoting sequential engagement of the IL-21 and IL-23 pathways. Nat Immunol. 2007;8(9):967–74. Epub 2007/06/22. doi: 10.1038/ni1488. PubMed PMID: 17581537.

55. McGeachy MJ, Cua DJ. The link between IL-23 and Th17 cell-mediated immune pathologies. Semin Immunol. 2007;19(6):372–6. Epub 2008/03/06. doi: 10.1016/j.smim.2007.10.012. PubMed PMID: 18319054.

56. Duerr RH, Taylor KD, Brant SR, Rioux JD, Silverberg MS, Daly MJ, et al. A genome-wide association study identifies IL23R as an inflammatory bowel disease gene. Science. 2006;314(5804):1461-3. doi: 1135245 [pii];10.1126/science.1135245 [doi].

57. Dubinsky MC, Wang D, Picornell Y, Wrobel I, Katzir L, Quiros A, et al. IL-23 receptor (IL-23R) gene protects against pediatric Crohn’s disease. Inflamm Bowel Dis. 2007;13(5):511–5. doi: 10.1002/ibd.20126 [doi].

58. Sandborn WJ, Gasink C, Gao LL, Blank MA, Johanns J, Guzzo C, et al. Ustekinumab induction and maintenance therapy in refractory Crohn’s disease. N Engl J Med. 2012;367(16):1519–28. doi: 10.1056/NEJMoa1203572 [doi].

59. Feagan BG, Sandborn WJ, D’Haens G, Panes J, Kaser A, Ferrante M, et al. Induction therapy with the selective interleukin-23 inhibitor risankizumab in patients with moderate-to-severe Crohn’s disease: a randomised, double-blind, placebo-controlled phase 2 study. Lancet. 2017;389(10080):1699–709. doi: 10.1016/S0140-6736(17)30570-6. PubMed PMID: 28411872.

60. Sands BE, Chen J, Feagan BG, Penney M, Rees WA, Danese S, et al. Efficacy and Safety of MEDI2070, an Antibody Against Interleukin 23, in Patients With Moderate to Severe Crohn’s Disease: A Phase 2a Study. Gastroenterology. 2017;153(1):77–86 e6. doi: 10.1053/j.gastro.2017.03.049. PubMed PMID: 28390867.

61. Almradi A, Hanzel J, Sedano R, Parker CE, Feagan BG, Ma C, et al. Clinical Trials of IL-12/IL-23 Inhibitors in Inflammatory Bowel Disease. Biodrugs. 2020;34(6):713–21. Epub 2020/10/27. doi: 10.1007/s40259-020-00451-w. PubMed PMID: 33105016.

62. Tsuda K, Yamanaka K, Kondo M, Matsubara K, Sasaki R, Tomimoto H, et al. Ustekinumab improves psoriasis without altering T cell cytokine production, differentiation, and T cell receptor repertoire diversity. PLoS One. 2012;7(12):e51819. Epub 2012/12/20. doi: 10.1371/journal.pone.0051819. PubMed PMID: 23251632; PubMed Central PMCID: PMCPMC3522598.

63. Jack C, Mashiko S, Arbour N, Bissonnette R, Sarfati M. Persistence of interleukin (IL)-17A+ T lymphocytes and IL-17A expression in treatment-resistant psoriatic plaques despite ustekinumab therapy. Br J Dermatol. 2017;177(1):267–70. Epub 2016/09/07. doi: 10.1111/bjd.15029. PubMed PMID: 27599204.

64. Leonardi CL, Kimball AB, Papp KA, Yeilding N, Guzzo C, Wang Y, et al. Efficacy and safety of ustekinumab, a human interleukin-12/23 monoclonal antibody, in patients with psoriasis: 76-week results from a randomised, double-blind, placebo-controlled trial (PHOENIX 1). Lancet. 2008;371(9625):1665–74. Epub 2008/05/20. doi: 10.1016/S0140-6736(08)60725-4. PubMed PMID: 18486739.

65. Papp KA, Langley RG, Lebwohl M, Krueger GG, Szapary P, Yeilding N, et al. Efficacy and safety of ustekinumab, a human interleukin-12/23 monoclonal antibody, in patients with psoriasis: 52-week results from a randomised, double-blind, placebo-controlled trial (PHOENIX 2). Lancet. 2008;371(9625):1675–84. Epub 2008/05/20. doi: 10.1016/S0140-6736(08)60726-6. PubMed PMID: 18486740.

66. Chiba A, Tajima R, Tomi C, Miyazaki Y, Yamamura T, Miyake S. Mucosal-associated invariant T cells promote inflammation and exacerbate disease in murine models of arthritis. Arthritis Rheum. 2012;64(1):153–61. doi: 10.1002/art.33314 [doi].

67. Guggino G, Di Liberto D, Lo Pizzo M, Saieva L, Alessandro R, Dieli F, et al. IL-17 polarization of MAIT cells is derived from the activation of two different pathways. Eur J Immunol. 2017;47(11):2002–3. Epub 2017/08/18. doi: 10.1002/eji.201747140. PubMed PMID: 28815578.

68. Wang H, Kjer-Nielsen L, Shi M, D’Souza C, Pediongco TJ, Cao H, et al. IL-23 costimulates antigen-specific MAIT cell activation and enables vaccination against bacterial infection. Sci Immunol. 2019;4(41). Epub 2019/11/17. doi: 10.1126/sciimmunol.aaw0402. PubMed PMID: 31732518.

69. Lord J, Chen J, Thirlby RC, Sherwood AM, Carlson CS. T-cell receptor sequencing reveals the clonal diversity and overlap of colonic effector and FOXP3+ T cells in ulcerative colitis. Inflamm Bowel Dis. 2015;21(1):19–30. Epub 2014/12/02. doi: 10.1097/MIB.0000000000000242. PubMed PMID: 25437819; PubMed Central PMCID: PMCPMC4526221.

70. Dash P, Wang GC, Thomas PG. Single-Cell Analysis of T-Cell Receptor alphabeta Repertoire. Methods Mol Biol. 2015;1343:181–97. doi: 10.1007/978-1-4939-2963-4_15. PubMed PMID: 26420718.

71. Lord JD, Shows DM. Thiopurine use associated with reduced B and natural killer cells in inflammatory bowel disease. World J Gastroenterol. 2017;23(18):3240–51. doi: 10.3748/wjg.v23.i18.3240. PubMed PMID: 28566883; PubMed Central PMCID: PMCPMC5434429.

72. Reantragoon R, Corbett AJ, Sakala IG, Gherardin NA, Furness JB, Chen Z, et al. Antigen-loaded MR1 tetramers define T cell receptor heterogeneity in mucosal-associated invariant T cells. J Exp Med. 2013;210(11):2305–20. doi: jem.20130958 [pii];10.1084/jem.20130958 [doi].

73. Lepore M, Kalinichenko A, Colone A, Paleja B, Singhal A, Tschumi A, et al. Parallel T-cell cloning and deep sequencing of human MAIT cells reveal stable oligoclonal TCRbeta repertoire. Nat Commun. 2014;5:3866. Epub 2014/05/17. doi: 10.1038/ncomms4866. PubMed PMID: 24832684.

74. Kim J, Chang DY, Lee HW, Lee H, Kim JH, Sung PS, et al. Innate-like Cytotoxic Function of Bystander-Activated CD8(+) T Cells Is Associated with Liver Injury in Acute Hepatitis A. Immunity. 2018;48(1):161–73 e5. Epub 20180102. doi: 10.1016/j.immuni.2017.11.025. PubMed PMID: 29305140.

75. Allez M, Skolnick BE, Wisniewska-Jarosinska M, Petryka R, Overgaard RV. Anti-NKG2D monoclonal antibody (NNC0142-0002) in active Crohn’s disease: a randomised controlled trial. Gut. 2017;66(11):1918–25. Epub 2016/08/05. doi: 10.1136/gutjnl-2016-311824. PubMed PMID: 27489241.

76. Cepek KL, Shaw SK, Parker CM, Russell GJ, Morrow JS, Rimm DL, et al. Adhesion between epithelial cells and T lymphocytes mediated by E-cadherin and the alpha E beta 7 integrin. Nature. 1994;372(6502):190-3. Epub 1994/11/10. doi: 10.1038/372190a0. PubMed PMID: 7969453.

77. Gold MC, McLaren JE, Reistetter JA, Smyk-Pearson S, Ladell K, Swarbrick GM, et al. MR1-restricted MAIT cells display ligand discrimination and pathogen selectivity through distinct T cell receptor usage. J Exp Med. 2014;211(8):1601–10. Epub 2014/07/23. doi: 10.1084/jem.20140507. PubMed PMID: 25049333; PubMed Central PMCID: PMCPMC4113934.

78. Standaert-Vitse A, Jouault T, Vandewalle P, Mille C, Seddik M, Sendid B, et al. Candida albicans is an immunogen for anti-Saccharomyces cerevisiae antibody markers of Crohn’s disease. Gastroenterology. 2006;130(6):1764–75. Epub 2006/05/16. doi: 10.1053/j.gastro.2006.02.009. PubMed PMID: 16697740.

79. McKenzie H, Main J, Pennington CR, Parratt D. Antibody to selected strains of Saccharomyces cerevisiae (baker’s and brewer’s yeast) and Candida albicans in Crohn’s disease. Gut. 1990;31(5):536–8. Epub 1990/05/01. doi: 10.1136/gut.31.5.536. PubMed PMID: 2190866; PubMed Central PMCID: PMCPMC1378569.

80. Forcione DG, Rosen MJ, Kisiel JB, Sands BE. Anti-Saccharomyces cerevisiae antibody (ASCA) positivity is associated with increased risk for early surgery in Crohn’s disease. Gut. 2004;53(8):1117–22. Epub 2004/07/13. doi: 10.1136/gut.2003.030734. PubMed PMID: 15247177; PubMed Central PMCID: PMCPMC1774147.

